# Contemporary burden of atrial fibrillation in mechanical thrombectomy stroke patients in the United States

**DOI:** 10.1101/2024.02.15.24302910

**Authors:** Fadar Oliver Otite, Smit D. Patel, Haydn Hoffman, Ehimen Aneni, Nnabuchi Anikpezie, Emmanuel Oladele Akano, Claribel Wee, Devin Burke, Karen Albright, Timothy Beutler, Julius Gene Latorre, Amit Singla, Nicholas Morris, Seemant Chaturvedi, Priyank Khandelwal

## Abstract

**Background:** How the prevalence of atrial fibrillation (AF) differs between various demographic subgroups of acute ischemic stroke (AIS) patients undergoing mechanical thrombectomy (MT) in the United States (US) is unknown. Data on whether AF prevalence in MT admissions changed over the last decade are sparse.

**Methods:** We conducted a serial cross-sectional study using all primary AIS discharges in the 2010-2020 National Inpatient Sample. Discharges with MT codes were identified (n=155,277) and the proportion with AF in various age, sex and racial subgroups were computed. We used multivariable-adjusted negative binomial regression to compare AF prevalence between demographic subgroups and Joinpoint regression to evaluate trends over time. Multivariable-adjusted generalized linear models were used to evaluate the association of AF with in-hospital outcomes.

**Results:** Across the study period, 45.0% of AIS discharges with MT had AF but marked disparity existed by age, sex and race. After multivariable adjustment, AF prevalence was 4% higher in women vs. men (prevalence rate ratio [PRR] 1.04, 95%CI 1.01-1.07), was lower in Black vs White (PRR 0.80, 95%CI 0.77-0.84) but higher in Asian compared to White discharges (PRR 1.11, 95%CI 1.05-1.18). Prevalence increased with age (PRR for ≥80 years vs 18-39 years: 5.23, 95%CI 4.28-6.39). Following joinpoint regression, prevalence increased by 3.2% (95%CI 1.3 to 5.2%) annually across the period 2010-2015 but declined by -2.2% (95%CI -2.9% to -1.4%) from 2015-2020. AF was associated with 27% lower odds of in-hospital mortality (Odds ratio 0.72, 95%CI 0.62-0.84) and 31% greater odds of routine home discharge (Odds ratio, 95%CI 1.17 to 1.47) compared to no AF.

**Conclusion:** AF prevalence in MT patients in the US is approximately twice that of the general AIS population but marked disparity exists by age, sex and race. AF Prevalence in MT increased from 2010-2015 but declined in the period 2015-2020.

**Clinical Perspective:** *What is new?:*

- Approximately 45% of mechanical thrombectomy acute ischemic stroke hospitalizations have comorbid atrial fibrillation (AF) and this proportion increases with age with 70% of patients >=80 years having AF.
- In contrast to all AIS patients, in the subset of AF patients undergoing MT, AF is associated with reduced in-hospital mortality and better odds of routine home discharge.

*What are the clinical implications?:*

- Given the post-stroke CHA_2_DS_2_-VASc score of at least 2, almost half of all MT patients in the US may be potential candidates for therapeutic anticoagulation.

## Introduction

Atrial fibrillation (AF) is the most common sustained arrhythmia^1^ and is associated with increased risk of acute ischemic stroke (AIS)^2^. Over 20% of all AIS admissions in the United States (US) have comorbid AF^3^, but prevalence estimates in AIS with large vessel occlusion (LVO) may likely be higher^4^. Whereas mechanical thrombectomy (MT) is now guideline-recommended therapy for AIS with LVO and its utilization has increased significantly in the US over the last decade^5^, how AF prevalence varies between various demographic subgroups of these MT-treated LVOs is not known. AF prevalence increases with age^6^ and may disproportionately affect men compared to women, although women >=80 years may be affected by AF more often^6,7^. Black people may also have lower prevalence of AF compared to White people^8^, but the age and/or sex groups of MT-treated LVOs in which these racial disparities in prevalence may be most prominent, are not currently known.

With aging of the population^9^, AF prevalence in MT-treated AIS over the last decade may have increased. However, advances in primary care and AF treatment over this time period, including more frequent utilization of any or direct oral anticoagulants in the US over the last decade,^10^ may have led to fewer AF patients presenting with LVO. Thus, it remains unknown how AF prevalence in MT patients has changed over time.

Furthermore, compared to non-AF AIS, AF-associated AIS have been shown to carry a more dismal outcome^11^. Infarct size tends to be larger and hemorrhagic transformation risk typically greater in AF-associated AIS compared to non-AF AIS^11^. However, whether outcomes differ between AF and non-AF patients in the subset of LVO patients undergoing MT remains very controversial^4,12^.

The primary aim of this study is to (1) Obtain age, sex and race specific prevalence of AF in LVO patients undergoing MT (2) Evaluate trends in the prevalence of AF in MT-treated LVOs in the US over the last decade (3) Compare in-hospital mortality and odds of routine home discharge between AF and non-AF associated AIS in the US.

## Methods

### Standard protocol approvals and data availability

This study was conducted using the National Inpatient Sample (NIS), one of the datasets from the Healthcare Cost and Utilization Project (HCUP). According to HCUP, utilization of the de-identified NIS does not require an Institutional Review Board review. The NIS is publicly available for direct purchase from HCUP. The authors are bound by data use agreement not to share HCUP data, but data analyses codes used in this study can be made readily available to interested parties on reasonable request.

We used data contained in the 2010–2020 NIS to conduct a serial cross-sectional study. The NIS is the largest publicly available inpatient care database in the US and consists of a 20% stratified random sample of all acute inpatient discharges in US hospitals. Further details on the NIS are available at hcup-us.ahrq.gov.

### Study population

We identified all adults (≥18 years) primary AIS discharges by querying the NIS using International Classification of Diseases, Ninth Revision (ICD-9) codes 433.X1, 434.XX and 436 before October 2015 and International Classification of Diseases, Tenth Revision (ICD-10) codes I63.XX afterward. These codes have been validated previously and found to be concordant with physician-diagnosed AIS in >90% of cases^13^. To minimize the risk of double counting hospitalizations, we excluded all discharges with length-of-stay of <24 hours and discharge to another short-term/acute hospital. All elective discharges were also excluded.

### Definition of covariates

Discharges with diagnostic codes for MT were defined using *ICD-9-CM* procedure code 39.74 and *ICD-10-CM* codes 03CG3ZZ, 03CG3Z6, 03CG3Z7, 03CG4Z6, 03CG4ZZ, 03CH3ZZ, 03CH3Z7, 03CJ3ZZ, 03CJ3Z7, 03CK3ZZ, 03CK3Z7, 03CL3ZZ, 03CL3Z7, 03CP3ZZ, 03CP3Z7, 03CQ3ZZ and 03CQ3Z7. AF was defined using ICD-9 CM codes 427.3x and ICD-10 codes in the range of I48.xx. These codes have been validated previously and shown to have very high accuracy for AF^14–17^. Hospitalizations with intravenous thrombolysis (IV-tPA) were defined using *ICD-9-CM* procedure code 99.10 or *ICD-10-CM* code 3E03317. We also classified discharges as those for IV-tPA if they had *ICD-9-CM* code V45.88 and *ICD-10-CM* code Z92.82, corresponding to IV-tPA administration within 24 hours at an outside facility.

Furthermore, we reviewed the discharge Medicare Severity Diagnosis Related Group codeand categorized admissions under the IV-tPA if they had Medicare Severity Diagnosis Related Group codes in the range of 061 to 063 (for “ischemic stroke with thrombolytic agents with or without comorbidity/conditions or major complication/comorbidity”). Admissions with secondary intracerebral hemorrhage were identified using secondary discharge codes in the range of 430, 431 or 432.9 (ICD-9) or ICD-10 codes I60-I62.

We used the “Elixhauser” add-on package in Stata to compute the Elixhauser score, a validated comorbidity score consisting of 31 comorbidities for all patients^18^. The National Institutes of Health Stroke Scale (NIHSS) was defined using *ICD-10* codes R29.7xx. Baseline CHA_2_DS_2_-VASc of all admissions were computed using components of the score (Table S1). Race and sex were determined using the HCUP variables “RACE” and “FEMALE”. Further descriptions of these variables are available at https://hcup-us.ahrq.gov/db/vars/race/nisnote.jsp and https://hcup-us.ahrq.gov/db/vars/female/nisnote.jsp

In-hospital all-cause mortality was defined using the HCUP variable “DIED”. We defined good outcome as routine home discharge, and this was defined using the HCUP variable “DISPUNIFORM”. In-hospital length of stay was determined using the HCUP variable LOS and total charges for each discharge determined using the variable “TOTCHG”.

### Statistical analysis

We summarized baseline characteristics of MT-treated AIS with and without AF using descriptive statistics. We computed the weighted prevalence of AF in MT admission subgroups categorized by age, sex and race. Statistical test of differences in prevalence between demographic subgroups was evaluated using the Pearson chi-square test. We further used negative binomial regression adjusted for differences in demographic factors, clinical and hospital level variables to compare prevalence rate of AF between demographic subgroups.

We also computed the annual prevalence of AF in MT-treated AIS and used joinpoint regression to evaluate trends in prevalence over time. Joinpoint uses a series of Monte-Carlo based simulations to identify points of change in trends (joinpoints) in a dataset. A regression line to the natural logarithm of the rates using calendar year as the regressor variable is then fitted to compute annualized percentage change (APC) for each identified trend.

We further used generalized linear models adjusted for demographic factors, stroke severity (NIHSS), Elixhauser score (comorbidity burden) and other hospitalization factors to evaluate differences in odds of in-hospital death between hospitalizations with prevalent AF and those without. Similar models were also used to evaluate the odds of secondary intracerebral hemorrhage or routine home discharge in hospitalizations with AF compared to those without.

Other generalized linear models were used to evaluate the association of AF with in-hospital length-of-stay or total charges. Independent variables in multivariable models were selected apriori based on their likely association with AF or AF detection or with hospitalization outcome. Because of the potentially strong confounding effect between stroke severity and these outcomes, we evaluated these outcomes only in admissions with available NIHSS. We however conducted additional analysis in all patients with and without this variable but excluding this variable from the multivariable model. We considered the weighting and clustering needed in the complex NIS survey data analysis. To account for changes in the NIS survey design in 2012, we used relevant NIS “TRENDWT” as recommended by HCUP. Because this is a descriptive analysis with no specific analysis tested, adjustment for multiple comparison was not necessary. A p-value of <0.05 was required for statistical significance in all analysis. All analysis were done by FOO using Stata 16 (College Station, Texas) and Joinpoint software version 4.8.0.1 (Bethesda, Maryland).

### Missing variables

The ICD-10 codes for NIHSS in administrative database was introduced in 2016 and was available in 64.9% of MT admissions from 2016-2020. Most other variables except race (5.4%) were missing in < 2.5% of admissions. Admissions with missing race were categorized into an unknown category. Admissions with missing information on other variables were imputed to the dominant category. Missing insurance data were categorized into the Medicare category if age ≥65 years and into the dominant category otherwise. For the NIHSS, multivariable models were conducted in all patients but not including the NIHSS and in additional models restricted only to patients with available scores.

## Results

Of 392,302,031 weighted hospital discharges in the US from 2010-2020, prevalence of comorbid AF in these discharges increased from 9.4% in 2010 to 13.8% in 2020 (Figure S1). Among the 5,190,148 primary AIS admissions in the US over this time, 3.0% (n=155,277) underwent MT. 45.0% of these MT admissions had comorbid AF. Baseline characteristics of MT admissions with and without AF are summarized in table 1. MT admissions with AF were relatively older and more likely to be white (Table 1). Baseline CHA_2_DS_2_-VASc scores, NIHSS and Elixhauser comorbidity scores were significantly higher in AF compared to non-AF admissions (table 1). The proportion of AF admissions with codes for IV-tPA (36.2%) and for mechanical ventilation (24.8%) were significantly lower compared to non-AF admissions (39.5% and 26.0% respectively, p-values <0.0001)

**Table 1.** Baseline characteristics of mechanical thrombectomy discharges in the United States from 2010-2020 according to atrial fibrillation status.

| **Variables** | **No atrial fibrillation** | **Atrial fibrillation** | **P-value** |
| --- | --- | --- | --- |
| Total Number | 17,128 | 13,989 | N/A |
| Weighted number, % | 85,469 (54.5) | 69,809 (45.5) | N/A |
| Female, % | 45.0 | 55.0 | <0.0001 |
| Age, years, mean (SE) | 63.4 (0.1) | 75.3 (0.1) | <0.0001 |
| **Age, %** |  |  | <0.0001 |
| 18-39 years | 6.5 | 0.8 |  |
| 40-59 years | 31.8 | 8.9 |  |
| 60-79 years | 47.1 | 48.4 |  |
| >=80 years | 14.7 | 41.9 |  |
| **Race, %** |  |  |  |
| White | 62.6 | 69.5 | <0.0001 |
| Black | 17.1 | 10.1 |  |
| Hispanic | 8.2 | 7.9 |  |
| Asian/Pacific Islander | 2.8 | 3.9 |  |
| Other/unknown | 9.3 | 8.6 |  |
| **Insurance status, %** |  |  | <0.0001 |
| Medicare | 48.3 | 76.3 |  |
| Medicaid | 13.3 | 5.3 |  |
| Private insurance | 29.5 | 14.4 |  |
| Self-pay | 5.7 | 2.2 |  |
| Other | 3.2 | 1.8 |  |
| **Clinical factors, %** |  |  |  |
| Hypertension | 74.5 | 85.4 | <0.0001 |
| Diabetes mellitus | 27.3 | 29.1 | 0.0013 |
| Dyslipidemia | 51.4 | 56.9 | <0.0001 |
| Coronary artery disease | 23.0 | 31.6 | <0.0001 |
| Congestive heart failure | 17.4 | 34.6 | <0.0001 |
| Dementia | 4.5 | 9.6 | <0.0001 |
| Intravenous thrombolysis | 39.5 | 36.2 | <0.0001 |
| Mechanical ventilation | 26.0 | 24.8 | 0.0214 |
| NIHSS, mean(SE) | 14.6 (0.1) | 16.3 (0.1) |  |
| Elixhauser comorbidity score | 4.3 (0.02) | 5.7 (0.02) | <0.0001 |
| CHADSVASc | 2.9 (0.01) | 4.1 (0.02) | <0.0001 |
| **Hospital characteristics, %** |  |  |  |
| Hospital region |  |  | <0.0001 |
| Northeast | 17.1 | 18.2 |  |
| Midwest | 21.5 | 21.3 |  |
| South | 40.9 | 36.8 |  |
| West | 20.4 | 23.6 |  |
| Hospital location/teaching status |  |  | 0.0136 |
| Rural | 0.3 | 0.2 |  |
| Urban non-teaching | 8.5 | 9.5 |  |
| Urban teaching | 91.2 | 90.3 |  |
| Length-of-stay, Days | 8.8 | 8.3 | <0.001 |
| Total Charges, \$ | 201,577 | 191,283 | <0.001 |
| In-hospital mortality | 13.2 | 13.0 | 0.4957 |
| Routine home discharge | 14.2 | 22.9 | <0.0001 |

N/A means not applicable.

### Age and sex differences in prevalence

AF prevalence in MT increased by a factor >1.5 fold with each 20-year increase in age, such that 70.0% of individuals ≥80 years old had comorbid AF (Figure 1). Overall AF prevalence was significantly greater in women (50.4%) compared to men (39.5%) (p<0.0001), but upon stratification by age, the age-specific prevalence was significantly higher in men in all age groups < 60 years, but greater in women compared to men in MT admissions ≥60 years (Figure 1).

**Figure 1.**
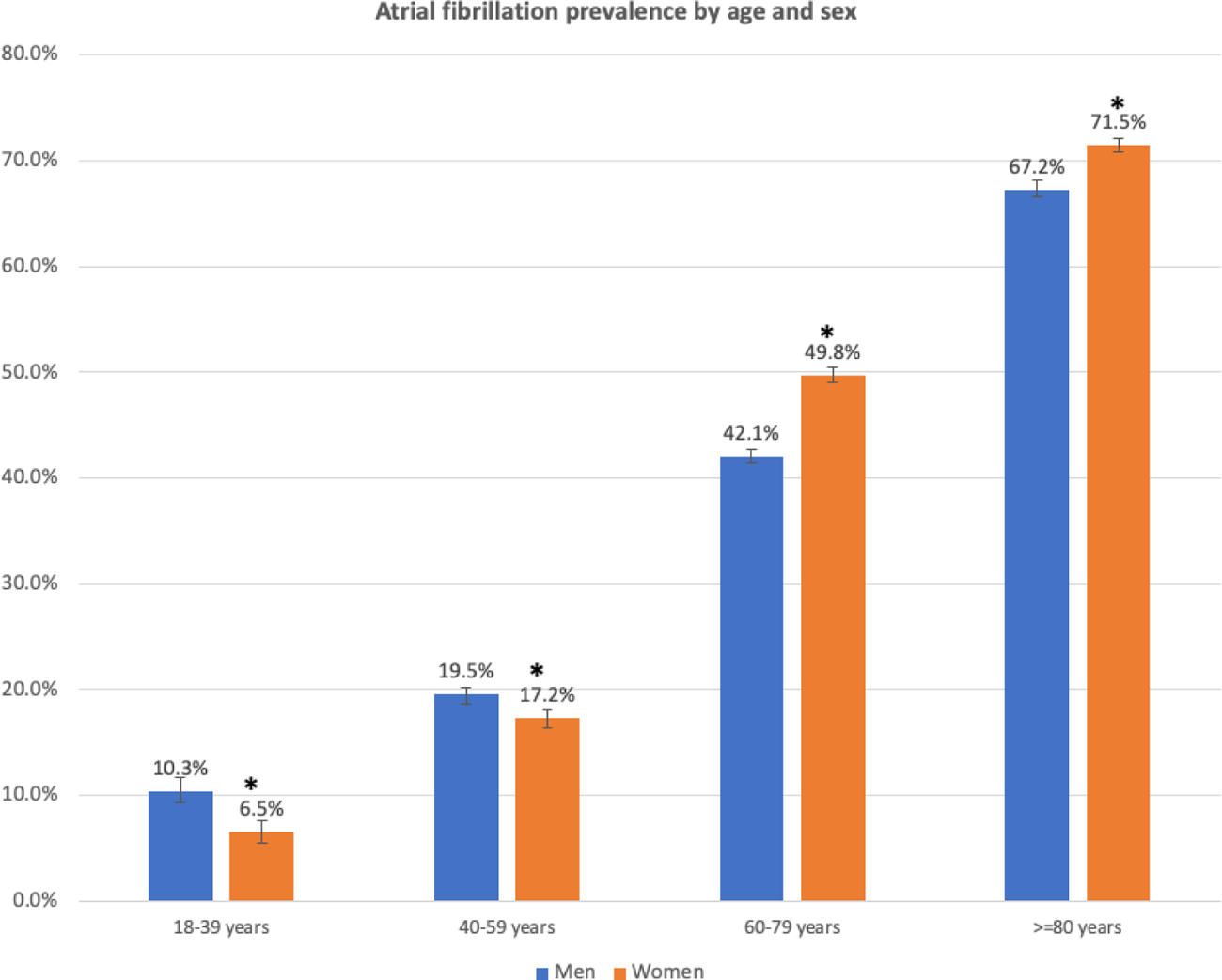
Prevalence of atrial fibrillation in mechanical thrombectomy discharges in the United States according to age and sex. *Indicates p-value <0.05 for comparison between men and women. Error bars represent standard error.

### Racial differences in prevalence

Unadjusted prevalence of AF was greater in admissions in White people (47.7%) compared to Black admissions (39.3%). This disparity in prevalence was driven by markedly higher prevalence in White admissions in people 60-79 years and ≥80 years old, as there was no significant difference in prevalence between Black admissions and White admissions in age groups below 60 years (Figure 2). In contrast, unadjusted AF prevalence in MT admissions in Asians was significantly greater compared to prevalence in White admissions, primarily due to higher AF prevalence in admissions in individuals 40-59 years and 60-79 years of age (Figure 2).

**Figure 2.**
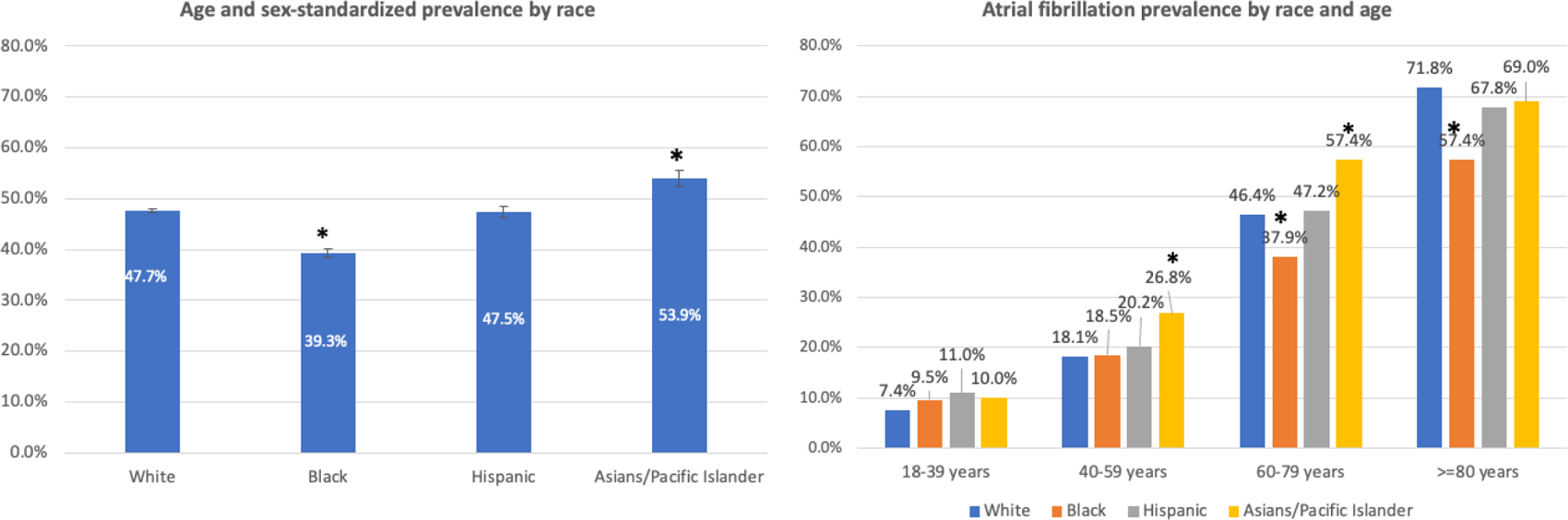
Prevalence of atrial fibrillation in mechanical thrombectomy discharges in the United States according to age and race. *Indicates p-value <0.05 for comparison of racial estimate to that of white admissions (in the same age group in age-stratified estimates). Absence of * indicates p-value for comparison > 0.05

### Multivariable-adjusted difference in prevalence between demographic subgroups

After multivariable adjustment for age, other demographic, clinical and hospital level factors, AF prevalence in MT admissions in Black people was 20% less compared to prevalence in White people (prevalence rate ratio [PRR] 0.80, 95%CI 0.76-0.84), but prevalence in Asian admissions was 11% greater compared to White admissions (PRR 1.11, 95%CI 1.05-1.18) (Table 2). Prevalence in women was 4% greater than prevalence in men (PRR 1.04, 95%CI 1.01 to 1.07) and increased with age. Prevalence of AF in ≥80-year-olds was more than 5-times the prevalence in young individuals 18-39 years (Table 2).

**Table 2.** Multivariable-adjusted comparison of the prevalence of atrial fibrillation in mechanical thrombectomy ischemic stroke discharges in the United States from 2010-2020.

| Variables | Prevalence ratio* | 95% CI | P-value |
| --- | --- | --- | --- |
| Race |  |  |  |
| Black vs white | 0.80 | 0.77 to 0.84 | <0.001 |
| Hispanic vs White | 0.97 | 0.92 to 1.01 | 0.176 |
| Asian/Pacific Islander vs White | 1.11 | 1.05 to 1.18 | <0.001 |
| Women vs men | 1.04 | 1.01 to 1.07 | 0.005 |
| Age Group |  |  |  |
| 40-59 years vs 18-39 years | 2.0 | 1.7 to 2.5 | <0.001 |
| 60-79 years vs 18-39 years | 3.96 | 3.25 to 4.83 | <0.001 |
| >=80 years vs 18-39 years | 5.23 | 4.28 to 6.39 | <0.001 |

\*Model further adjusted for income, hospital region, hospital location/teaching status, smoking, dyslipidemia , Elixhauser comorbidity score, dyslipidemia, dementia, smoking status, intravenous thrombolysis, CHA<sub>2</sub>DS<sub>2</sub>-VASc score and year.

### Trends in Prevalence over time

After joinpoint regression, AF prevalence increased by 3.2% annually over the period 2010-2015, (APC 3.2%, 95%CI 1.3% to 5.2%, p=0.006) but declined by approximately 2% across the period 2015-2020 (APC -2.2%, 95%CI -2.9% to -1.4%, p=0.001) (Figure 3). Most of the initial increase in AF prevalence and subsequent decline across the study period occurred in admissions in patients 60-79 years and ≥80-year-olds, as the prevalence in younger patients did not change significantly across the study period (Figure 3). Age and sex-adjusted prevalence also declined in Black and White discharges across the period 2015-2020 (Figure S2).

**Figure 3.**
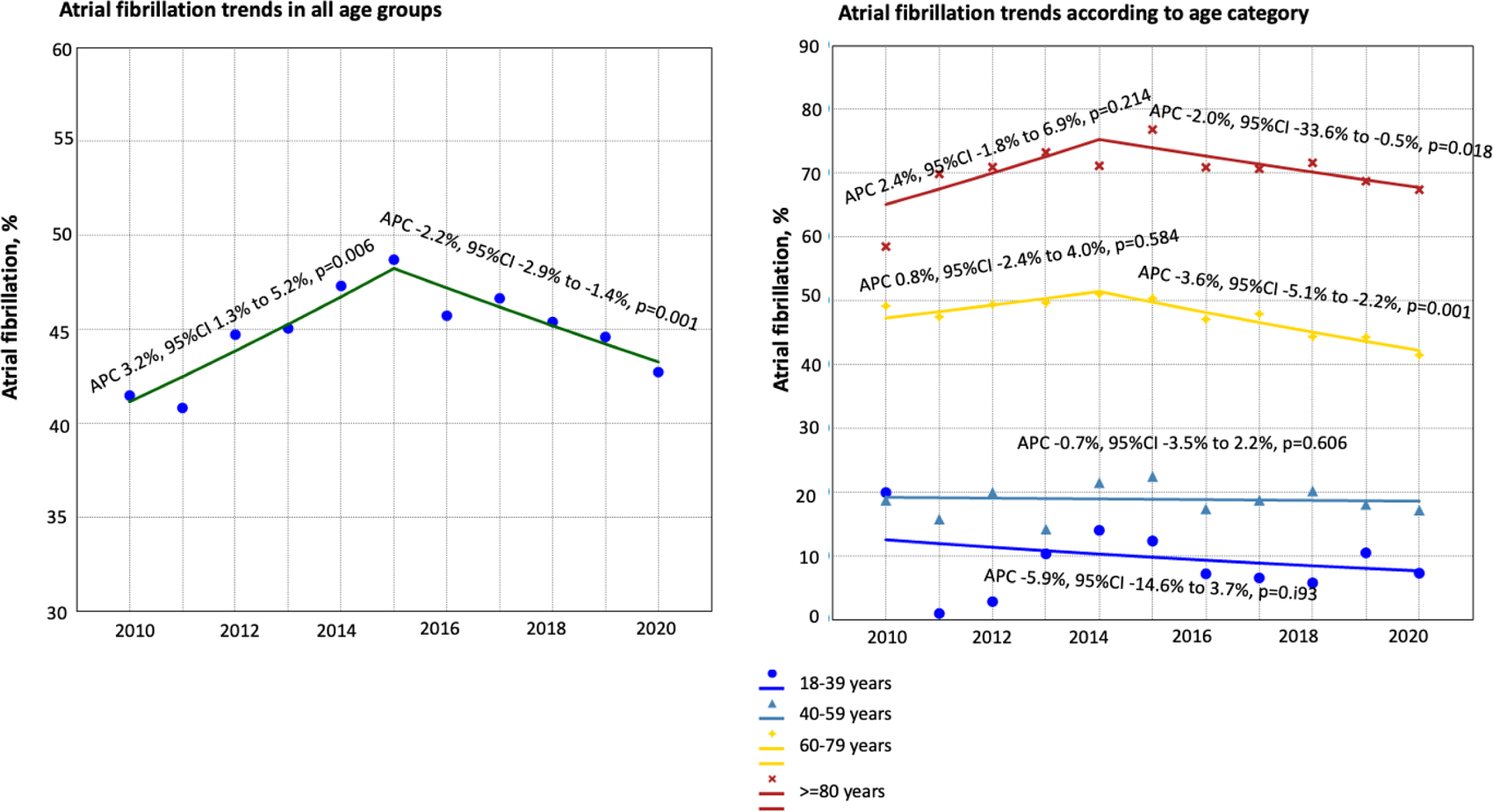
Trends in the prevalence of atrial fibrillation in mechanical thrombectomy discharges in the United States from 2010-2020 APC represents annualized percentage change.

### Multivariable association of AF with secondary intracerebral hemorrhage, in-hospital mortality and odds of routine home discharge

After multivariable adjustment for differences in age, stroke severity (NIHSS) and other demographic and hospital level factors, AF was associated with reduced odds of in-hospital mortality and increased odds of routine home discharge in all admissions and in the subset of admissions with available NIHSS. For example among admissions with non-missing NIHSS, MT admissions with AF had approximately 25% lower odds of in-hospital mortality (OR 0.73, 95%CI 0.63 to 0.85) compared to those without AF. Admissions with AF also had approximately 30% greater odds of routine home discharge (OR 1.32, 95%CI 1.17 to 1.48) compared to those without AF in admissions with available NIHSS. Odds of secondary intracerebral hemorrhage did not differ between discharges with and without comorbid AF (OR 1.05, [95%CI 0.96 to 1.16]) but mean length-of-stay was shorter and mean total hospital charges at least $13,000 less in AF compared to non-AF discharges (Table 3).

**Table 3.** Association of atrial fibrillation with odds of in-hospital outcomes following mechanical thrombectomy.

| **Variables** | **Model estimates\*** | **95% CI** | **P-value** |
| --- | --- | --- | --- |
| **All admissions** |  |  |  |
| **Intracerebral hemorrhage** | 1.02 | 0.95 to 1.08 | 0.637 |
| **Length-of-stay, days** | -0.66 | -0.83 to -0.50 | <0.001 |
| **Total charges, \$** | \$-13,484.72 | -16,476.48 to -10,492.96 | <0.001 |
| **In-hospital mortality** | 0.80 | 0.73 to 0.88 | <0.001 |
| **Routine home discharge** | 1.33 | 1.22 to 1.45 | <0.001 |
| **Admissions with available National Institute of Health Stroke Scale** |  |  |  |
| **Intracerebral hemorrhage** | 1.05 | 0.94 to 1.26 | 0.322 |
| **Length-of-stay, days** | -0.73 | -0.94 to -0.51 | <0.001 |
| **Total charges, \$** | -14,367.29 | -18,814.45 to -9,920.14 | <0.001 |
| **In-hospital mortality** | 0.72 | 0.62 to 0.84 | <0.001 |
| **Routine home discharge** | 1.31 | 1.17 to 1.47 | <0.001 |

\*Estimates represent odds ratio for dichotomous variables (intracerebral hemorrhage, in-hospital mortality and routine home discharge) and beta coefficient for continuous variables.
All models adjusted for age, sex, race insurance status, intravenous thrombolysis, National Institute of Health Stroke Scale, smoking status, dementia, Do-Not-Resuscitate, coronary artery disease, mechanical ventilation, coma/stupor, dyslipidemia, Elixhauser comorbidity score, hospital region, hospital location/teaching status, year of discharge.

## Discussion

In this contemporary NIS analysis, we found that 45.0% of MT admissions in the US had comorbid AF which is approximately twice the prevalence in the general AIS population. However, marked disparity in prevalence existed by age, sex and race. After multivariable adjustment, prevalence increased with age and was marginally higher in women compared to men. Prevalence was approximately 20% higher in White compared to Black admissions, but 10% lower in White when compared to Asian/Pacific Islander admissions. Overall AF prevalence increased in the first half of the decade from 2010-2015 but declined significantly in the latter half from 2015-2020 mainly in patients ≥60 years old. MT admissions with comorbid AF had approximately 25% lower odds of all-cause in-hospital mortality and 30% greater odds of routine home discharge compared to those without AF.

Accurate understanding of demographic variations and trends in AF prevalence is critical for discerning changes in the epidemiologic characteristics and outcome of LVO AIS. Multiple prior MT studies have reported crude AF prevalence in MT, but large-scale studies systematically evaluating its prevalence in various demographic subgroups are hitherto lacking. Amongst all participants enrolled in MT trials contained in the HERMES collaboration meta-analysis, the prevalence of AF was 33%^19^. However, the clinical characteristics of these highly selected clinical trials patients may not be generalizable to all MT patients in the real world. In another meta-analysis of studies evaluating outcomes in MT patients with AF, the prevalence of AF in the ten included studies ranged from 20-55%^4^. In this study we provide new information highlighting AF prevalence in various age, sex and racial groups that are generalizable to all MT patients in the US.

Although it has been well established that AF prevalence increases with age^6^, the exceptionally high prevalence in patients ≥80 years (70.0%) in this study is notable and implies that clinicians taking care of very elderly MT patients may need to consider a more rigorous search for underlying AF in this age group when the source of a suspected embolic stroke remains unidentified. Further subgroup analysis of our data revealed sex differences in AF prevalence with increasing age. Whereas AF was more prevalent in men in admissions <60 years of age, prevalence was greater in women in admissions ≥60 years. AF in women carry a higher AIS risk compared to men^20^ but this excess risk may not be very apparent in individuals <75 years of age^21^. Other major causes of LVO AIS such as extracranial carotid atherosclerosis^22^ may be more prominent etiologies of AIS in men compared to women. All these factors may potentially contribute to the higher AF prevalence noted in the very elderly women with MT compared to similar aged men. In addition, the underutilization of anticoagulants in women may lead to presentation with an AF-related AIS.

Our finding of higher prevalence of AF in White compared to Black admissions is consistent with previously known racial differences in community-wide AF prevalence. However, the higher prevalence of AF in Asian/Pacific Islander admissions compared to White admissions undergoing MT is surprising partly because AF is less prevalent in the general population^8^ and in AIS in Asian compared to White people^23^. Moreover, ICAD may account for up to half of all AIS in Asians^24^ and so one would expect ICAD to account for a disproportionately greater proportion of LVO in Asian admissions in this study. Whether Asians/Pacific Islanders with AF in the US are more prone to LVO stroke compared to other races or how racial differences in detected AF may be contributing to these differences remains unknown.^25^ It is also possible that the relatively higher AF in White compared to most minority racial/ethnic groups may be biased by racial disparity in MT access.

In this study we report on the prevalence of AF but are unable to ascertain an etiological relationship between AF and AIS. We are also unable to differentiate between AF present before admission or AF detected after stroke (AFDAS). AFDAS may represent a subset of AF that may carry lower AIS recurrent risk^26,27^. Nevertheless, given that the CHA_2_DS_2_-VASc scores of all these AF patients will be no less than 2 after stroke, our findings imply that almost half of all MT patients in the US may be potential candidates for therapeutic anticoagulation based on current guidelines^28^. In the very elderly patients ≥80 years, this proportion may even be higher: every 7-in-10.

The rising prevalence of AF in MT in the period 2010-2015 is consistent with data from our prior study and those of others reporting increased prevalence of AF in all AIS in the US over the decade from 2003-2014^3,29^. However, the decline in AF prevalence noted over the period 2015-2020 in this study is unexpected and the underlying reason for this finding is not very apparent. This decline was despite observed increase in AF prevalence in all hospitalizations over the study period. It is very likely that concerted public health efforts towards ensuring appropriate primary stroke prevention in individuals with AF through more frequent use of therapeutic anticoagulation may be playing a major role in this decline. More widespread usage of NOACs and other measures for secondary stroke prevention in AF patients such as left atrial appendage closure may likely be playing a role. However, there is still considerable room for improvement in use of DOACs for stroke prevention in AF^30^. Future studies targeted at understanding these changes are needed.

Prior studies evaluating the association of AF with outcomes following MT have yielded conflicting results. Whereas the HERMES collaboration meta-analysis of MT randomized trials found no difference in odds of Modified Rankin Scale of 0-2 at 90 days between AF and non-AF patients^31^, another meta-analysis of 11 randomized trials reported a positive association between AF and good outcome^12^. A more recent meta-analysis of ten retrospective observational studies reported 47% greater odds of 90-day mortality in AF compared to non-AF patients^4^ but this study did not account for difference in age between AF and non-AF patients^4^, a major confounder of its association with mortality. All of these studies are very small in comparison to the current study and recruited patients mainly from large academic centers that may not be reflective of the total MT population in the US. That AF is associated with increased mortality and poorer functional outcome compared to other AIS subtypes has been established in multiple cohorts^32–34^. Our data however suggests that in the subset of AIS patients undergoing MT, AF may be associated with lower odds of in-hospital mortality and better odds of routine home discharge. In-hospital LOS and corresponding charges of hospital stay are also significantly less in AF compared to non-AF MT discharges. The exact underlying reasons for this possible better outcome in AF MT admissions still requires further evaluation in prospective registries but it is possible that faster MT procedural times in AF patients, an important determinant of MT outcomes may potentially be responsible for this . In one recent analysis of 4,169 MT patients contained in the Stroke Thrombectomy and Aneurysm Registry (STAR) procedural times were faster, MT passes were fewer, and rates of first pass success were higher in AF compared to non-AF admissions^35^. Moreover, some forms of non-AF LVO such as those due to underlying ICAD carry high risk of MT failure, defined as thrombectomy in Cerebral infarction grade less than 2^36^ and MT failure may be associated with larger AIS core and worse functional outcome compared to AF-associated strokes.^36^

This study has additional limitations. Although the ICD-codes used for AIS and AF have been shown to have high accuracy, we cannot exclude coding errors. Our study of trends in AF prevalence over time is based on the implicit assumption that coding practices remained unchanged over time. However, improved coding is likely to have led to increased AF documentation and prevalence over time, yet we observe a decline over the period 2015-2020. The number of codes per encounter retained in administrative datasets such as the NIS has increased over time^15^, and this is expected to also lead to increased prevalence of comorbidities not the decline we are noting for AF. In fact, AF in all hospitalizations in the NIS also increased over the years 2010-2020. Our study period bridges that between ICD-9 and ICD-10, and this may be a potential source of coding error. The general algorithm to categorize AF did not change from ICD-9 to ICD-10^15^ so no major difference in AF prevalence is likely to be attributable to changes in ICD coding. Our evaluation of secondary ICH should be viewed with caution as the sensitivity of ICD-10 codes for this complication is still unknown.

This study likely underestimates the true burden of AF in MT as a significant proportion of paroxysmal AF continues to be diagnosed beyond AIS hospitalization. We have no specific information on the location of LVO, timing from presentation to recanalization or the degree of recanalization, all of which may potentially be associated with AIS outcome. Methods for obtaining race/ethnicity data in the NIS is not standardized across hospitals or states and is not always based on the gold standard of self-report. We have no information on non-binary gender expressions because these are set to missing in the NIS.

## Conclusion

Of all hospitalizations in AIS patients undergoing MT in the United States over the decade from 2010-2020, 45.0% had comorbid atrial fibrillation and this proportion increased with age, varied with sex and was disproportionately higher in Asians compared to White and in White compared to Black discharges. Prevalence increased over time from 2010-2015 and declined from 2015-2020. Future studies are needed to understand the reasons underlying this decline.

## Data Availability

All data used in this study can be purchased directly from the Healthcare Cost and Utilization Project. The authors are bound by data use agreeement not to share HCUP data

## Nonstandard Abbreviations and Acronyms

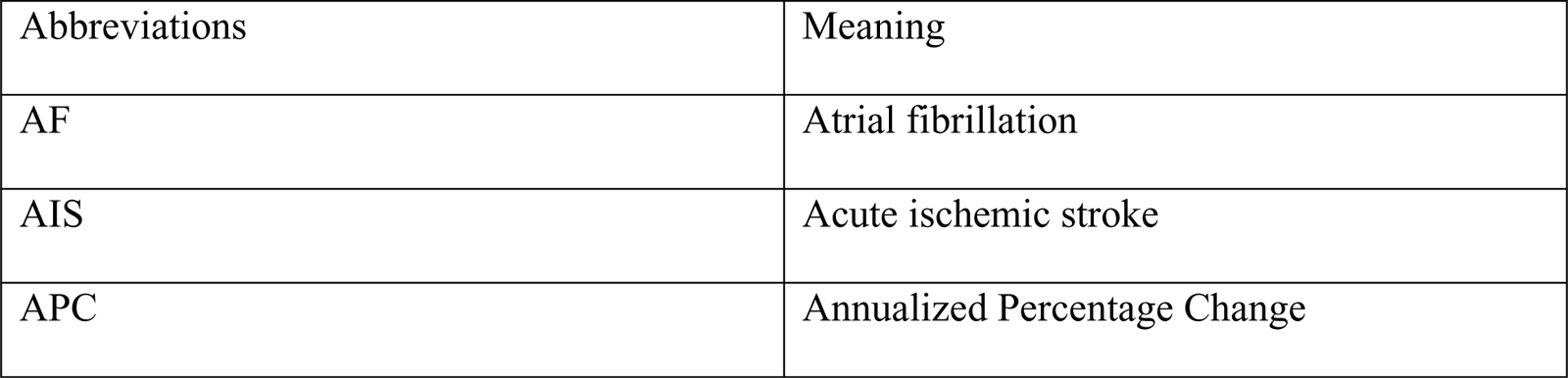

| Abbreviations | Meaning |
| --- | --- |
| AF | Atrial fibrillation |
| AIS | Acute ischemic stroke |
| APC | Annualized Percentage Change |
| HCUP | Healthcare Cost and Utilization Project |
| ICD | International Classification of Diseases |
| LVO | Large vessel occlusion |
| NIS | National Inpatient Sample |
| MT | Mechanical thrombectomy |
| US | United States |

## Acknowledgement

none.

